# Maternal characteristics and pregnancy outcomes during the global pandemic Covid-19 in indonesian tertiary referral hospital

**DOI:** 10.1101/2023.02.27.23286514

**Authors:** Sofia Al Farizi, Dewi Setyowati, Azra Fauziyah Azyanti, Dyah Ayu Fatmaningrum, Rahayu Kusuma Candrakirana

## Abstract

**Aim:** Analyse differences in intervention and pregnancy outcomes characteristics in obstetric patients with a diagnosis of COVID-19 and non-COVID-19 at one of the Indonesia tertiary referral hospital in East Java.

**Design:** This was cross sectional study.

**Methods:** This study was performed 694 obstetric patients, the data for these patients were obtained from the hospital medical records Sampling was used simple random. This study used Mann-Whitney test to analyse the differences between the variables.

**Results:** There was a significant difference in the Length Of Stay (LOS), LOS of COVID-19 patients tends to be longer than that of non-COVID-19 patients. More than half of the patients gave birth by caesarean delivery, 83 for COVID-19 and 283 for Non COVID-19. Some of the most common complications among COVID-19 patients were maternal infectious and parasitic diseases (1.3% vs 0.0%), abnormalities of forces of labour (12.3% vs 9.6%), complication of puerperium (0.6 % vs 0.0%). 40.9% COVID-19 patient suffered Acute Respiratory Distress Syndrome (ARDS). COVID-19 infection had no significant effect on pregnancy outcomes.

**Conclusion:** Several interventions need to be re-evaluated, such as cesarean delivery in COVID-19 and non-COVID-19 patients. The health-care delivery system must also be re-evaluated, and the tiered referral system must be strengthened.

## Introduction

At the end of 2019, Coronavirus disease 2019 (COVID-19) infected Wuhan, China, and spread to several countries worldwide. Furthermore, World Health Organization (WHO) declared a COVID-19 pandemic in March, 2020 (1). This virus first entered Indonesia on March 2, 2020, after two positive cases were discovered. On April 9, 2020, COVID-19 cases began to spread in 34 provinces in Indonesia, including East Java (2). Hospitals in East Java are experiencing problems with health services because they are constrained by the lack of infrastructure, the availability of isolation rooms, and personal protective equipment (PPE) (3). As one of the referral hospitals in East Java experienced a density of patients during the COVID-19 explosion. Because of the large number of health workers who have contracted the COVID-19 virus, the number of health workers available to provide services is very limited, causing services to run inefficiently (4). The referral system in Indonesia so far also cannot run during the COVID-19 pandemic. Coordination between facilities and efforts to prevent primary health care is not running optimally (5).More research is needed to determine the impact of COVID-19 on mothers and babies. According to a retrospective review from Wuhan, China, pregnant women’s clinical characteristics are the same as those of non-pregnant women. Intrauterine transmission has also not been demonstrated (6). According to a UK study, COVID-19 has no significant negative impact to pregnant women, and the need for respiratory support is minimal (7). COVID-19 does not worsen the women’s condition and is not proven to cause transmission to the baby (8). Several researchers in Indonesia are taking an interest in this case. Previously, research at a secondary referral hospital in Surabaya, East Java, found that clinical manifestations and laboratory test results were similar in COVID-19 and non-COVID-19 patients. However, this study demonstrates that the risk of death in COVID-19 patients is high (5). Another study conducted at a hospital in Surakarta, Central Java, found that pregnant women infected with COVID-19 did not exhibit severe signs and symptoms (9).

This study aimed to analyse the characteristics of intervention and pregnancy outcomes in obstetric (maternal) patients in COVID-19 and non-COVID-19 patients in one of Indonesian tertiary referral hospital.

## Methods

### Design

This study was cross-sectional that utilized secondary data from medical records in one of the tertiary hospitals in East Java, Indonesia in 2020. The hospital was selected because it was one of the largest hospitals in Indonesia, so the number of patients with Covid-19 was found.

### Samples

The required sample size was calculated using simple random sampling. The sample size was estimated by Slovin’s formula. The population of this study were pregnant, labour and postpartum women, with as many as 1265 (N), 8 pregnant women, 391 labour women and 301 postpartum women. The used margin of error was 0,05 (e).

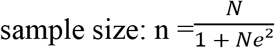

As a result, we decided to include 700 patients in this study. Because, in simple random sampling, the strong representativeness of the target population can increase with a large sample size (10) The sampling method was a simple random technique with several inclusion criteria such as obstetric patients (pregnancy, labour and postpartum), available medical record data, and all required information was complete. While the exclusion criteria were not obstetric cases, such as gynecological cases. Data collection was carried out from May 30, until August 1, 2021. The dataset was compiled from hospital patient records from March, 03, 2020 until February, 28, 2021.

### Data Collection

The medical record system in this hospital used electronic medical records and was equipped with documents to add data. Since this hospital was a teaching hospital, the accuracy and validity of the data in the medical records were actually maintained.

This study compared several variables in COVID-19 and non-COVID-19 women, including maternal demographic data (age, educational attainment, occupation and residence distribution), hospitalization characteristics (hospitalization admission procedure, delivery method, length of stay, diagnostic), and maternal outputs. In the maternal demographic data, we compared regions into four groups; Surabaya and Sidoarjo are two areas in East Java Province which was have big population. The categorization of diagnoses in this study was based on the ICD-10. Beside obstetric diagnosis, we also included several diagnoses related to COVID-19, such as acute respiratory distress syndrome, renal failure, and cardiac events.

### Data Analysis

In this study, general descriptive statistics such as frequency distributions were used. After that we compared the differences in several variables in the two groups. Mann Whitney analysis was used to compare the two research variables with p-value <0.05. SPSS ver. 25 (IBM; Armonk, NY, USA) was used to analyze the data.

### Ethical Statement

This research was approved by the Health Research Ethics Commission of RSUD Dr. Soetomo with No. 0495/LOE/301/.4.2/VI/2021.

## Results

The study included 694 pregnant, labour and postpartum women in total (154 mothers with COVID-19 and 540 mothers without COVID-19).

### The Maternal Demographic Characteristics

The maternal demographic characteristics who involved in this study showed in Table 1. Several variables showed the same trend in both groups, and there was no significant difference in both. Respondents in this study were dominated by women aged 21-25 years and 26-30 years of COVID-19 and non-COVID-19 patients, respectively. COVID-19 patients aged 21 to 25 years was 33.1%, and as many as 25.3% are aged 26-30 years. The same data was also shown by non-COVID-19 patients, as many as 24.8% of mothers aged 21-25 years and 31.5% aged 26-30 years. There were no significant differences in each age group in women with COVID-19 and non-COVID-19. The same data was indicated by maternal education attainment, and there was no significant difference between the two groups. Almost half of COVID-19 (50.6%) and non-COVID-19 patients (56.7%) have a complete vocation degree education level. This trend was also shown by mother occupation; most of the occupations in both groups are housewives, 57.8% of women suffering from COVID-19 are housewives, and 60.6% of women who are not COVID-19 patients are housewives. However, a different phenomenon was shown in the group of patients suffering from COVID-19. College students in this group were more in quantity than in the group of patients who did not suffer from COVID-19. From the operational definition we got in the hospital’s medical record, this college student is a resident student.

**Table 1.**
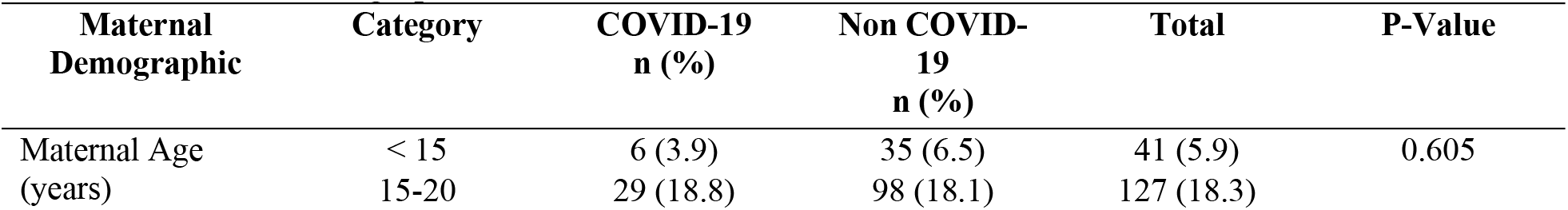
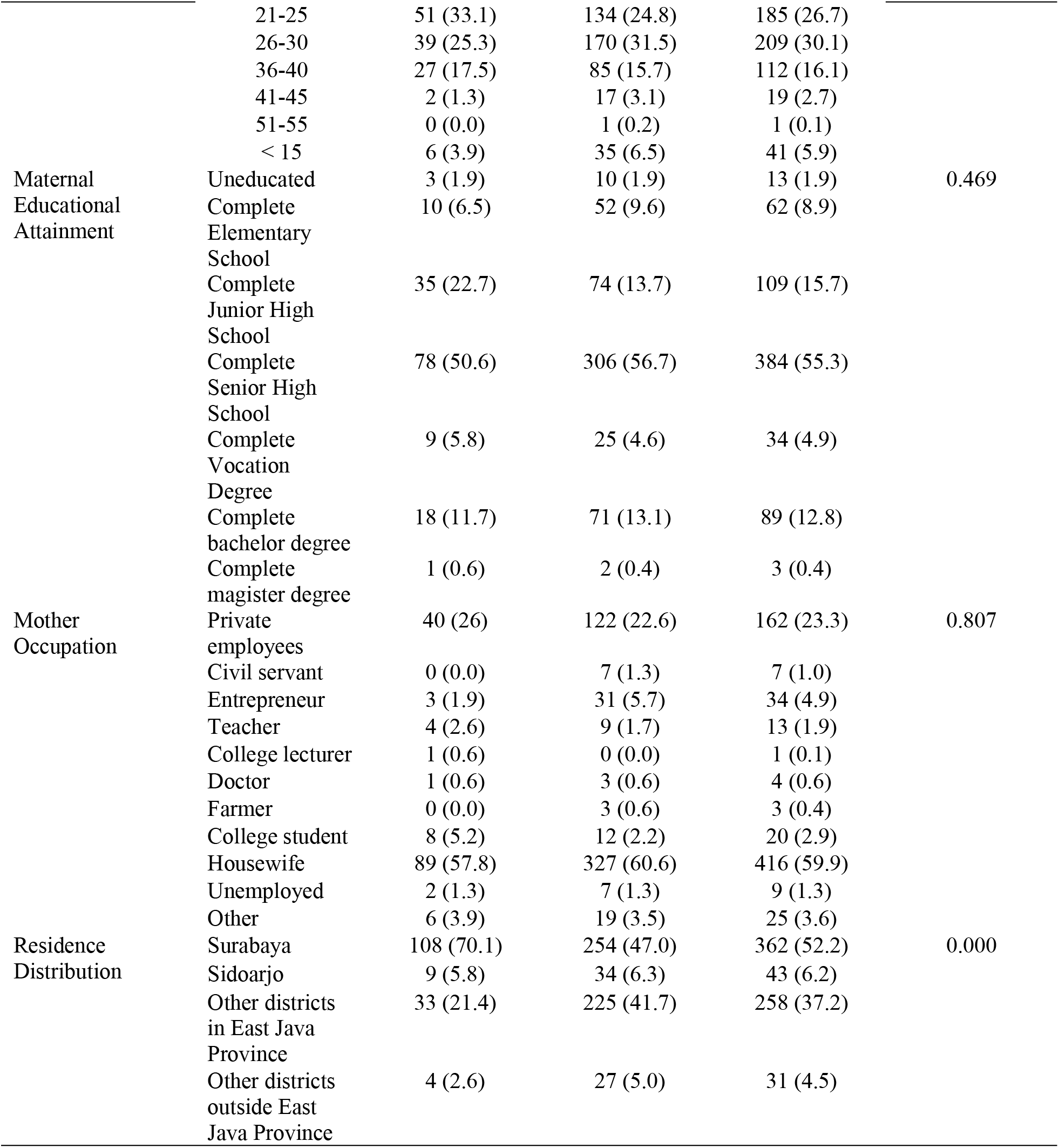
Maternal Demographic Characteristics

Residence distribution in the two groups shows a significantly different percentage. COVID-19 patients are dominated by women from Surabaya and Sidoarjo. 70.1% of COVID-19 patients came from the Surabaya area, while for non-COVID-19 patients, only 47.0%. Patients from other districts in East Java Province were actually more in the non-COVID-19 patient group, which was 41.7%. Patients from other areas outside of East Java were also more in the non-COVID-19 patient group, which was 5.0%.

### The Hospitalization Characteristics

Table 2 explained the hospitalization characteristic. There was a significant difference in the hospital admission procedure between the two groups. In patients without COVID-19, the referral process was still going well, 15.2% of patients were outpatient referrals. This is different from COVID-19 patients, only 5.2% were outpatient referrals. Most of the COVID-19 patients were emergency installation referrals (92.9%), while 1.9% were patients without referrals/inpatients. Indeed, most of the patients during the COVID-19 pandemic were dominated by emergency installation referrals.

**Table 2.**
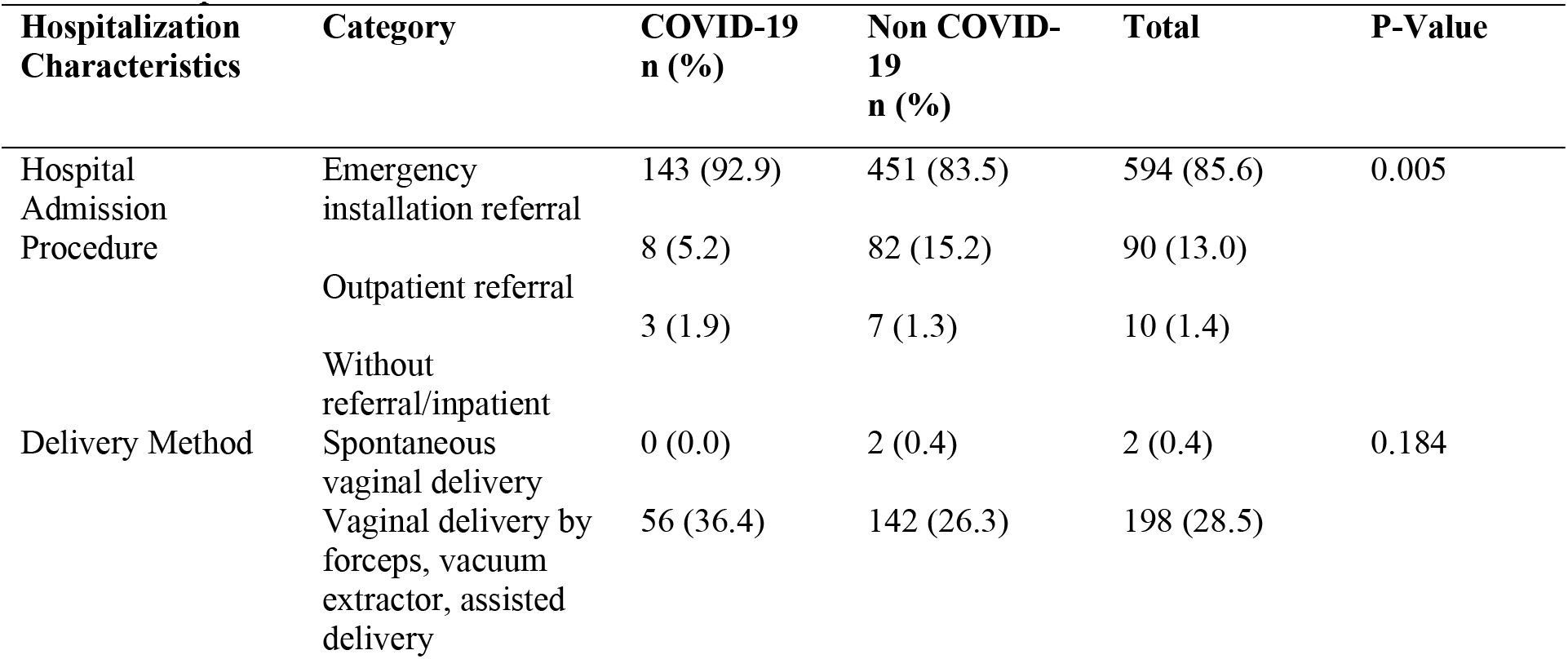
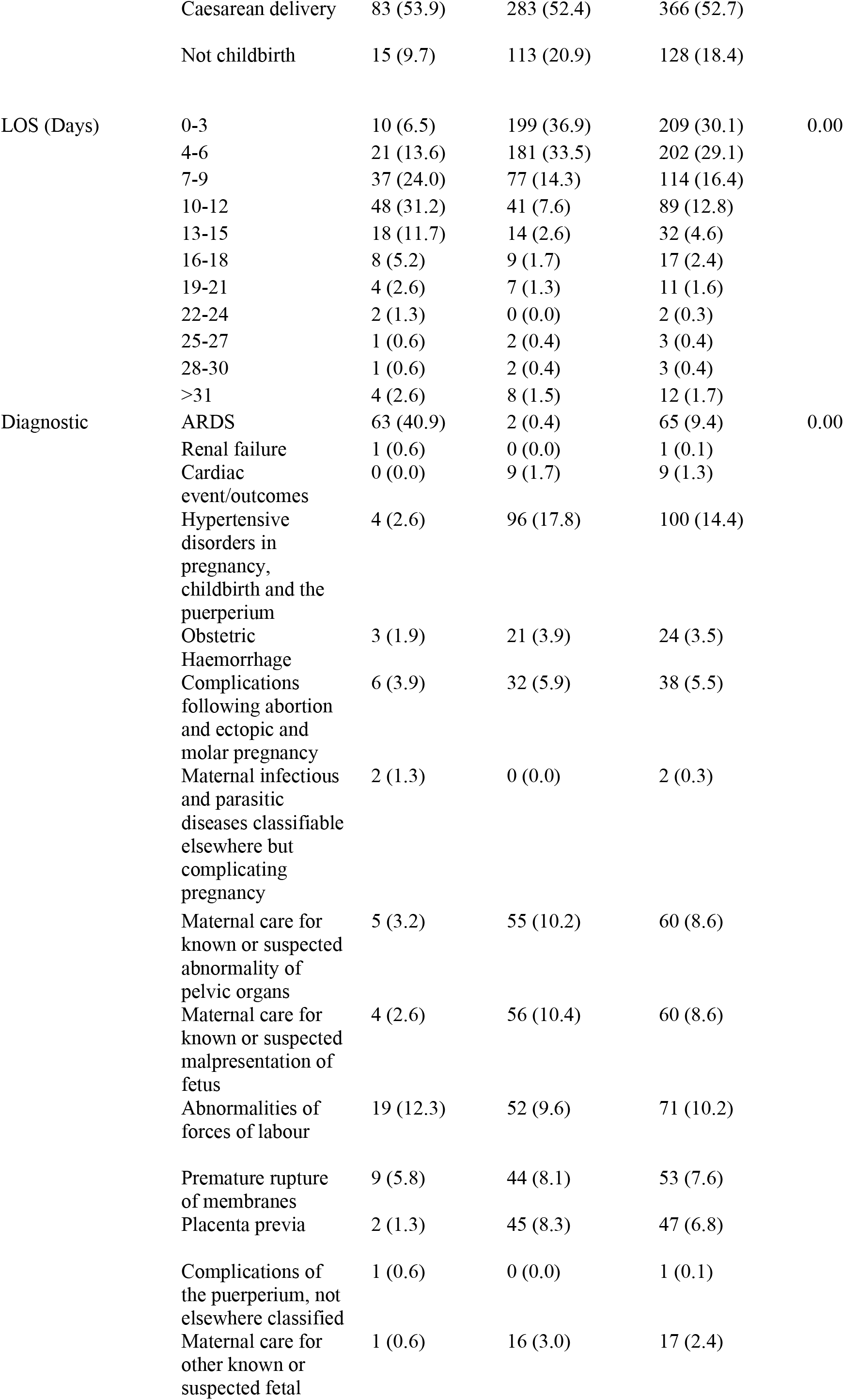
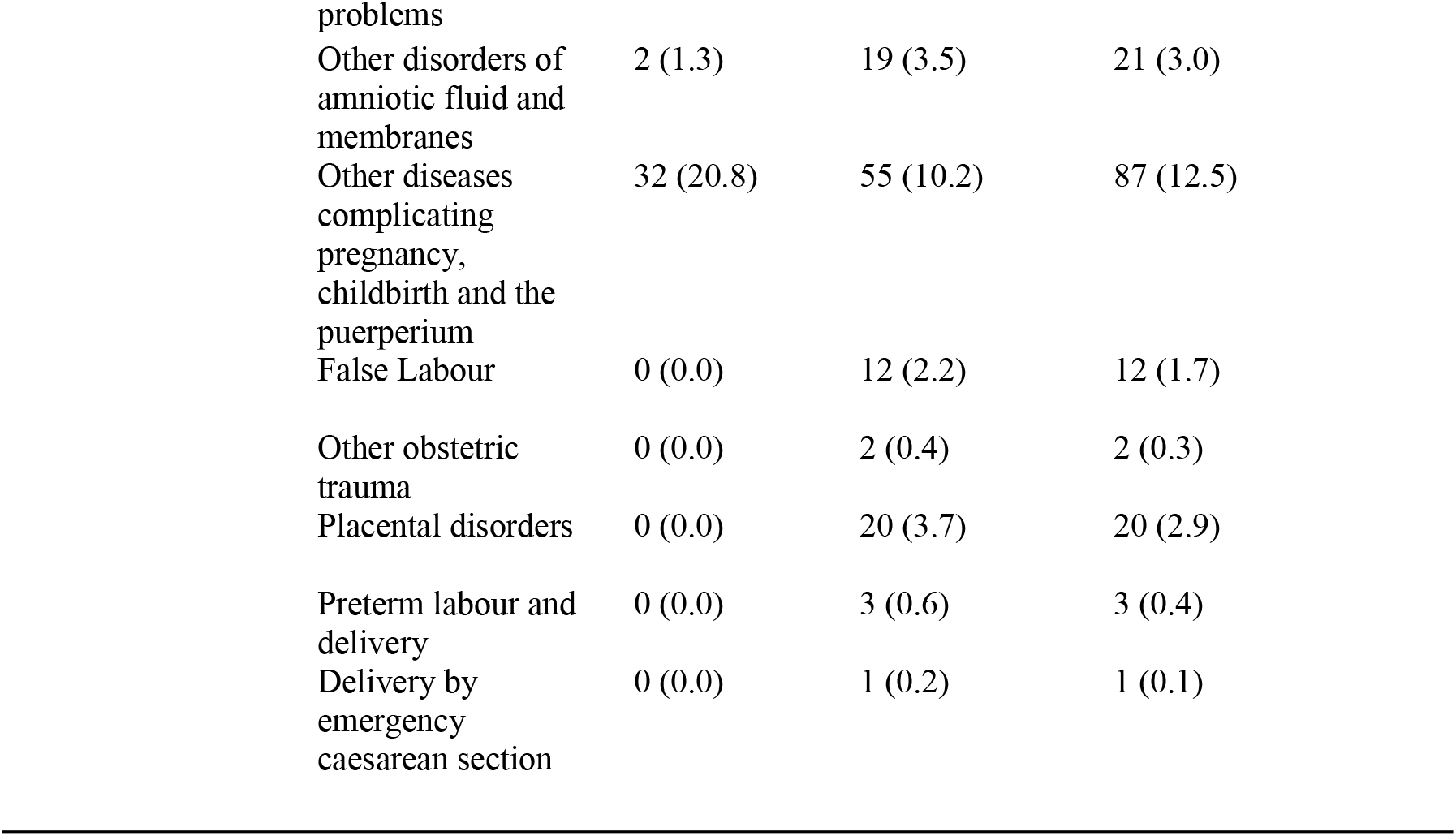
Hospitalization Characteristic

Delivery by caesarean delivery dominated in both groups, 53.9% of COVID-19 patients and 26.3% of non-COVID-19 patients. There was no significant difference in labour characteristics between the two groups. Vaginal delivery by forceps, vacuum extractor, and assisted delivery also showed almost the same numbers in the group, 36.4% in COVID-19 patients and 26.3% in non-COVID-19 patients. Meanwhile, spontaneous vaginal delivery was only available in non-COVID-19 patients, which was 0.4%. LOS in the two groups was significantly different, and COVID-19 patients tended to have a longer LOS when compared to non-COVID-19 patients. 31.2% of COVID-19 patients had a LOS of 10-12 days, while most non-COVID-19 patients (36.9%) had a shorter LOS of 0-3 days.

The diagnosis in the two groups had significantly different characteristics. Most of the COVID-19 patients had a diagnosis of ARDS (40.9%). Another obstetric diagnosis that exists in COVID-19 patients is abnormalities of forces of labour, and this complication is greater in COVID-19 patients than in non-COVID-19 patients (12.3% vs 9.6%). Most of the non-COVID-19 patients were diagnosed hypertensive disorders in pregnancy, childbirth and puerperium (17.8%). As for the case of death, the mortality of COVID-19 patients (4.5%) was more significant than non-COVID-19 patients (3.3%), but this difference was not significant.

### Maternal Outcomes

The maternal outcomes show in Table 3. There was no significant difference in maternal mortality (p = 0.47). However, the percentage of maternal mortality in COVID-19 patients tends to be higher than in non-COVID-19 patients (4.5% vs 3.3%).

**Table 3.** Maternal Outcomes

| Maternal Output | COVID-19<br>n (%) | Non COVID-19<br>n (%) | Total | p-value |
| --- | --- | --- | --- | --- |
| Death | 7 (4.5) | 18 (3.3) | 25 (3.6) | 0.447 |
| Not Death | 147 (95.5) | 522 (96.7) | 669 (96.4) |  |

## Discussion

The tiered referral system for maternal health services cannot run optimally during pandemic COVID-19. As a tertiary hospital, this hospital should be a patient referral centre in East Java. However, this was not seen in pregnant, maternity and postpartum patients during the COVID-19 pandemic. Patients from Surabaya and Sidoarjo dominate the treatment facilities at this hospital. In fact, 70.1% of COVID-19 patients came from Surabaya. Emergency installation referral became the dominant hospital admission procedure in both groups. The characteristics of the hospital admission procedure in the two groups were significantly different. One of the studies stated that most patients came to the obstetric emergency room for delivery purposes, so it will be challenging to find patients who have COVID-19 symptoms (11). COVID-19 patients without referral/inpatient are higher than non-COVID-19 patients. This condition can be an illustration that the referral system that Indonesia has implemented is not working well (12). The referral system during the COVID-19 pandemic is still showing a weak point. In reality, 76% of village health posts and 41% of home visits were closed. Primary health services cannot run optimally during the COVID-19 pandemic, such as family planning, immunization, and other routine maternal and child healthcare services. Impaired mobility due to travel restrictions is one of the factors for this. In addition to the fear of officers against transmission, primary and secondary health services do not run optimally (5,12).

More than half of COVID-19 patients (53.9%) gave birth by caesarean delivery method. Previous research also stated that there was no significant difference in the mode of delivery in -19 patients who had symptoms or not (11). Other referral hospitals also showed the same trend, that caesarean delivery dominated in COVID-19 patients (3). At the beginning of the pandemic, several referral hospitals in Indonesia did recommend caesarean delivery for COVID-19 patients, aiming to reduce the risk of transmission to health workers early in the pandemic (3,11). Some hypotheses suggest that the increase in caesarean delivery is due to regulatory changes to adapt to the COVID-19 pandemic (13). The National Obstetrics Gynaecology Association (POGI) recommends caesarean deliveries to COVID-19 patients (14). Vaginal delivery is considered to increase contact between patients and health workers because it requires longer observation, besides vaginal delivery contributes droplets to health workers when entering the 2nd stage of labour (3,15). Indeed, several countries have also made COVID-19 a medical indication for caesarean delivery interventions so that this method of delivery dominates (15). Several meta-analyses and systematic reviews also state that more than half of COVID-19 patients give birth by caesarean method (8,16). Overall, caesarean delivery did dominate during the COVID-19 pandemic, 52.7% of deliveries to mothers were caesarean. When viewed from the study results, caesarean delivery also dominated non-COVID-19 maternity mothers (52.4%). China is a country that has the same trend, caesarean delivery is the dominant medical intervention for women who are in lockdown (17). Although this action is medically justified, the relationship between caesarean delivery and neonatal maternal mortality is only 20% (18). The World Health Organization (WHO) also recommends that the limit of caesarean delivery does not exceed 15-20% (19).

COVID-19 patients tend to have significantly longer LOS when compared to non-COVID-19 patients. Most patients with COVID-19 had LOS for 10-12 days (31.2%), while 36.9% of non-COVID-19 patients had LOS for 0-3 days. In fact, prolonged length of stay occurs if the duration of treatment is three days after birth. So it can be concluded that COVID-19 patients tend to experience a prolonged length of stay compared to non-COVID-19 patients (20). Some countries show different phenomena, LOS during the pandemic decreased compared to before the pandemic, and LOS for COVID-19 patients tended to be short (20,21). However, it should be considered that the clinical degree of a disease can also indicate a prolonged LOS (22). Moreover, if the patient comes to the hospital in an already severe condition. Considering the data states that the entry procedure for COVID-19 patients is dominated by emergencies.

There was no significant difference in the incidence of maternal mortality in COVID-19 and non-COVID-19 patients. However, some diagnoses were found to be significantly higher in COVID-19 patients. First, maternal infectious and parasitic diseases classifiable elsewhere but complicating pregnancy (1.3% vs 0.0%), tuberculosis is a specific complication of this diagnosis. Pregnant women who have a history of tuberculosis have a greater risk of suffering from Severe Acute Respiratory Syndrome Coronavirus 2 (SARS-CoV-2), and the output obtained will be more labour intensive when compared to women who do not have this history (23). However, we did not get information on how tuberculosis travels in COVID-19 patients, so we cannot provide an analysis of whether tuberculosis appeared before they were diagnosed with COVID-19 or after. Furthermore, abnormalities of forces of labour in COVID-19 patients are also higher than in non-COVID-19 patients (12.3% vs 9.6%). In the ICD category, these complications are related to labour progress. Some complications such as inadequate contraction, uterine inertia, and hypertonic are categorized in this diagnosis. The high number of complications that affect labour progress can be a medical indication for cesarean delivery. Looking back at the recommendations made by POGI, obstacles in the progress of labour can be an indication for a cesarean delivery so that contact between health workers and patients can be minimized (14). Lastly, the diagnosis of complications of the puerperium, not elsewhere classified (0.6% vs 0.0%) was found only in patients with COVID-19. More specifically, disruption of caesarean section wound is a complication that occurs in this diagnosis. Certainly, this also correlates with the high percentage of cesarean deliveries in COVID-19 patients, so the possibility of complications due to this procedure is also higher.

### Limitation of This Study

This research was conducted only through secondary data, there has been no in-depth study through direct observation of respondents. In addition, this study does not capture the full picture of maternal health during the pandemic in Indonesia, which is about 2 years.

## Conclusion

This study illustrates no significant difference in the percentage of maternal deaths in COVID-19 and non-COVID-19 patients. However, several other output indicators show different things. The LOS of COVID-19 patients has been shown to be longer than that of non-COVID-19 patients, and the ARDS complications that can aggravate the mother’s condition are also relatively high. Another thing that needs to be a concern is the high rate of caesarean delivery during the COVID-19 pandemic, both for COVID-19 and non-COVID-19 patients. Also, the referral process is not running properly, so there is a risk of accumulating patients in the ER because emergency referrals dominate.

The health service system in Indonesia should be prepared for a crisis such as a pandemic. The government must intensify health service innovation, such as remote technology-based services, to improve this. Likewise, health workers, such as midwives, should already have breakthroughs and capabilities related to health service innovation. This, of course, will affect the quality of services provided.

## Data Availability

Yes, we will provide all the required data

